# Association between biomarkers of environmental enteric dysfunction and growth and neurodevelopment in Guatemalan infants

**DOI:** 10.1101/2024.11.10.24317056

**Authors:** Amy K. Connery, Diva M. Calvimontes, Filemon Bucardo, Daniel Olson, Alison M. Colbert, Luther Bartelt, Edwin J. Asturias, Molly M. Lamb, Sylvia Becker-Dreps

**Author notes:** Co-Senior Authors (Molly M. Lamb, Sylvia Becker-Dreps).

## Abstract

**BACKGROUND:** There is growing evidence that environmental enteric dysfunction (EED) is a driver of poor growth and neurodevelopment (ND) in early childhood. To further investigate this, we measured the associations between biomarkers reflecting various domains of EED and growth and ND in Guatemalan infants. METHODS. A subset of 114 cohort infants were randomly selected for inclusion from a 2017-2019 population-based cohort study of 499 young infants in rural southwest Guatemala. Growth and neurodevelopmental performance using the Mullen Scales of Early Learning (MSEL) were assessed at a household visit around 12 months of age. Serum samples collected at the visit were analyzed for concentrations of α-1 acid glycoprotein (AGP), glucagon-like peptide-2 (GLP-2), and anti-flagellin IgA (Anti-FliC IgA), Multivariable regression analyses adjusted for relevant confounders were conducted to define associations between these EED biomarkers and length-for-age z-score (LAZ) and neurodevelopmental performance. We planned *a priori* to conduct analyses with and without excluding infants who had acute infectious disease symptoms (fever, cough, vomiting, diarrhea) at the 12-month visit.

**RESULTS:** No significant association between biomarkers representing different domains of EED and LAZ or MSEL scores at 12-14 months of age was found. However, removing children with acute infectious symptoms uncovered an association between Anti-FliC IgA and MSEL. Specifically, an increase in 10ng/L in Anti-FliC IgA concentration was associated in a decrease in the MSEL Early Learning Composite (ELC) raw score of 3.2 points, which equates to approximately a 9-point decrease in the ELC standard score.

**CONCLUSIONS:** In this study, having increased levels of Anti-FliC IgA was associated with lower ND in the first year of life and may represent an important risk to long-term health and development.

**AUTHOR SUMMARY:** There is growing evidence that a key driver of stunting and poor neurodevelopment in low-resource settings is environmental enteric dysfunction. Environmental enteric dysfunction is a subclinical condition of the small intestine that may include chronic mucosal inflammation and/or increased intestinal permeability; it is common in children experiencing repeated infections with fecal-oral pathogens. In this preliminary study, we examined the link between biomarkers of environmental enteric dysfunction and neurodevelopment in 114 infants. After excluding children with acute infectious symptoms (which could transiently modify biomarker levels), we found a strong link between one marker of EED and poor neurodevelopment, independent of child growth. Further research is warranted to explore the associations between EED and ND to guide future interventions to reduce these risks.

## INTRODUCTION

Among global health inequities, disparities in child growth and neurodevelopment (ND) are perhaps the most devastating. Estimates suggest that almost half of children living in low- and middle-income countries (LMICs) are at risk for poor ND based on the prevalence of stunting(1) and that the gap in ND widens between these children and their healthy peers with the cumulative exposures of living in poverty.(2–4) There is also clear evidence that poor ND in early childhood can have lasting effects and can impact both educational achievement(5–7) and long-term human potential.(7–10)

Poor ND is associated with stunting, a manifestation of chronic undernutrition.(11–13) ND is especially vulnerable during infancy, a time of rapid growth and intricate development of the neurological system.(12,14) However, decades of nutritional interventions have not resulted in substantial improvements in growth and ND, suggesting that the problem is not simply one of a deficiency of calories or nutrients.(15–17) Therefore, recent efforts have focused on identifying other potential causal factors. There is now growing evidence that environmental enteric dysfunction (EED)—a subclinical small intestinal condition that may consist of chronic mucosal inflammation, villous blunting, increased intestinal permeability, growth hormone resistance, and/or dysbiosis of the gut microbiome—may be a key driver of chronic malnutrition and poor ND.(18–21) EED is thought to be caused by recurrent infections with fecal-oral pathogens from poor sanitation and water quality, and exposure to domestic animals.(22,23) The result of these changes is a reduced capacity for nutrient absorption leading to chronic malnutrition, impaired enteric immunity, and systemic inflammation.(16,24)

The gold standard for the diagnosis of EED is intestinal biopsy, which is not ethical to perform in observational studies of children and is also not feasible in many LMIC settings. Therefore, most studies of EED assess its various domains —intestinal permeability, health of the intestinal epithelium, and chronic inflammation— through the measurement of biomarkers. Recent studies from Pakistan, Tanzania, and Uganda have linked levels of certain EED biomarkers to child’s faltering growth,(25–27) a study from Tanzania linked EED biomarkers to poor ND(28), while two studies from Bangladesh have linked biomarkers of systemic inflammation to poor ND.(29,30) Given the consequences of poor growth and ND on overall child health and lifelong educational and career attainment, additional studies are needed to firmly establish this association, identify which domains of EED are most harmful, and plan targeted interventional studies. In addition, studies from more diverse settings are needed to better understand whether this association is universal across low-resource settings. The goal of this exploratory study was to understand the associations between biomarkers of EED and growth and ND in a birth cohort from a rural region Guatemala, known to have a high burden of poor linear growth and poor neurodevelopmental outcomes.(31,32)

## Methods

### Study Design and Clinical Assessments

From June 2017 to July 2019, we conducted a prospective cohort natural history study (“parent study”) of the incidence and sequelae of postnatally-acquired ZIKV infection in infants and young children at the Center for Human Development research and clinic site in southwest Guatemala.(33) The site encompasses primarily rural communities with approximately 25,000 residents and is located approximately 30 km from the border with Chiapas, Mexico. These communities are monolingual Spanish-speaking. The population suffers from high rates of food insecurity and child undernutrition, diarrheal disease, maternal depression, and maternal and child morbidity and mortality.(31,33–35)

We enrolled 499 infants aged 0-3 months and their mothers. Per parent study design,(33) there were no exclusion criteria other than living outside the study catchment area and having the appropriate age. Demographics, prenatal history, and birth information were collected at enrollment, including maternal education level on a 4-point scale (no schooling, primary, secondary, or university/postgraduate education), and housing material (cement vs. wood, aluminum, or other). Breastfeeding duration, age at introduction to liquids and complementary foods, and anthropometric data (weight, length, head circumference) were collected at enrollment and at 3-month intervals. Length-for-age z scores (LAZ) were calculated to determine stunting, defined as >2 standard deviations (SDs) below the mean on World Health Organization (WHO)-defined growth curves.(36) Following enrollment, households were visited weekly by trained study nurses, and caregivers were asked to report the presence or absence of each of the following symptoms in the infant in the last 7 days preceding their surveillance visit: fever, cough, vomiting/diarrhea (≥3 loose stools/day), rash, conjunctivitis (non-purulent/hyperemic), arthralgia, myalgia, or peri-articular edema. The weekly household visits were continued for 12 months.

Neurodevelopmental assessments were conducted at enrollment, and at approximately 6 and 12 months of age. Data from the 12-month follow-up assessment was used in the current study. Neurodevelopmental testing included the Mullen Scales of Early Learning (MSEL), following an extensive validation process at the site and with rigorous administration procedures, as previously described.(37–40) The MSEL is comprised of five subscales: Gross and Fine Motor, Expressive and Receptive Language, and Visual Reception. The scores from four of the scales (excluding Gross Motor) are summed to create the Early Learning Composite (ELC) Score. Given the lack of local, population-based norms, raw scores adjusted for age were used in all analyses instead of standard scores, which is best practice for translated and adapted tests.(41) This also helped us to detect more subtle differences among this group of children with multiple shared neurodevelopmental risk factors.

In this additional analysis of biobanked serum samples from the parent study, we randomly selected 150 participating children who had no evidence of Zika virus infection, and for whom we had complete outcome data at the final (12 month) study visit. One hundred and fourteen of these participants had a serum sample with sufficient volume to be analyzed for concentrations of three biomarkers of EED.

### Laboratory Analysis

Serum biomarkers associated with systemic inflammation, enterocyte repair, and intestinal permeability were selected to capture different pathological domains of EED. Human α1-acid glycoprotein (AGP) is an acute-phase serum protein normally produced by the liver at concentrations ranging from 50 to 130 mg/dL. In response to inflammation and infection, the serum level can increase more than two-fold as compared with levels in healthy individuals.(42–44) AGP serum concentrations were measured by the enzyme linked immunosorbent assay (ELISA) manufactured by R&D Systems, Inc (Quantikine® ELISA, Minneapolis, USA) following manufacturer’s instructions. In brief, 50 μL of serum or standard dilutions were added to microwells pre-coated with AGP monoclonal antibody (mAb). Optical density (OD, 450nm) of the ELISA reactions were measured in the BioTek Epoch 2 Microplate Spectrophotometer. Concentrations were determined in GraphPad (Version 10.0.0 for Windows, Boston, USA) by interpolating individual OD reading into sigmoidal 4PL model. Glucagon like peptide-2 (GLP-2) is normally produced in the enteroendocrine L cells from the mucosa of the distal small intestine and throughout the large intestine and is considered a marker of gastrointestinal repair.(45) GLP-2 acts to increase intestinal epithelial absorptive capacity and mucosal growth by activating enteric neurons.(46) GLP-2 levels are inversely related to nutrient absorption.(47) GLP-2 serum concentrations were measured by ELISA (Millipore Sigma, Darmstadt, Germany) following manufacturer’s instructions. In brief, 50 μL of serum were added to the wells pre-coated with a polyclonal rabbit anti-GLP-2 antibody. OD readings and GLP concentrations were determined as described above. Anti-flagellin IgA (Anti-FliC IgA) levels measure the host’s systemic exposure to bacterial components.(27,48) As there is a high concentration of bacterial flagellin in the intestinal lumen, Anti-FliC IgA can be a biomarker for gut barrier function.(48,49) Hence children with higher blood concentrations of these immunoglobulins are thought to have had greater exposure to gut bacteria as a consequence of impaired mucosal barriers.(16) Anti-FliC IgA concentrations in serum were measured by ELISA (SunLong Biotech Co. LTD, Hangzhou, China) following manufacturer’s instructions. In brief, 10μl of serum diluted 1:5 was added to the well pre-coated with flagellin antigen. The serum concentration of anti-flagellin IgA was determined as described above.

### Statistical Analysis

Descriptive statistics were used to characterize the study population, and histograms of the distribution of the EED biomarkers were created. Spearman’s correlation coefficients were used to estimate the correlations between concentrations of the three EED biomarkers. Bivariate and multivariable linear regression models were then used to test associations between each EED biomarker (AGP, GLP-2, and Anti-FliC IgA), and MSEL ELC score, Gross Motor (GM) subscale score and length-for-age z-score (LAZ) at the 12-month visit. First, separate bivariate analyses testing the association between each of the three EED biomarkers (AGP, GLP-2, and Anti-FliC IgA) with each of the three outcomes (ELC, GM, and LAZ), adjusted for infant’s age at the 12-month visit, were conducted. After identification of potentially relevant confounders via testing of the association between each potential confounder and both the exposures of interest and outcomes of interest in separate models, we selected all potential confounder variables that showed association (p< 0.10) with any of the exposures and/or outcomes to be included in the multivariate analysis. Final models were adjusted for infant’s age at 12-month visit, sex, maternal education level, housing material, and age at introduction to solid foods (Table 2). Finally, we were concerned that acute infectious illnesses may cause transient changes in EED biomarker levels that might mask an underlying association. Therefore, we decided *a priori* to perform the multivariate analysis separately for infants who did not have acute infectious disease symptoms (cough, diarrhea, vomiting, fever) at time of the 12-month visit with blood sampling (N = 63). All analyses were conducted using SAS version 9.4 (SAS Institute, Cary, NC).

### Ethical approvals

The parent study protocol was reviewed and approved by the Institutional Review Board (IRB) at Baylor College of Medicine, the Colorado Multiple Institutional Review, the National Ethics Committee of the Guatemalan Ministry of Public Health and the local Community Advisory Board in Trifinio, Guatemala. The mother of each child (or in the case of a minor mother, the child’s mother and the mother’s guardian) provided written informed consent for their child’s participation in the parent study and for future unspecified research using biobanked specimens. This additional analysis was approved by the IRB of the University of North Carolina at Chapel Hill.

## Results

The randomly selected sub-cohort of 114 infants was 39% female, mean LAZ was −1.56 (SD 0.94), 74% lived in a house made of cement blocks (a sign of higher socioeconomic status; SES), and the majority of mothers (58%) did not have an education beyond primary school (Table 1). In comparison, the parent study participants were 48% female, mean LAZ was −1.53 (SD 1.0), 78% lived in a house made of cement blocks, and 63% had a mother with a primary school education. The mean concentrations of the three EED biomarkers tested, AGP, GLP-2, and Anti-FliC IgA, were 654.93 µg/mL, 3.76 ng/mL, and 10.71 ng/L, respectively. The distributions of the concentrations of the three biomarkers are shown in Figure 1. In the correlation analysis, we found weak positive correlations between the concentrations of AGP and GLP-2 (ρ=0.32, p=0.0006) and between AGP and Anti-FliC IgA (ρ=0.23, p=0.01), as shown in Table 2.

**Figure 1.**
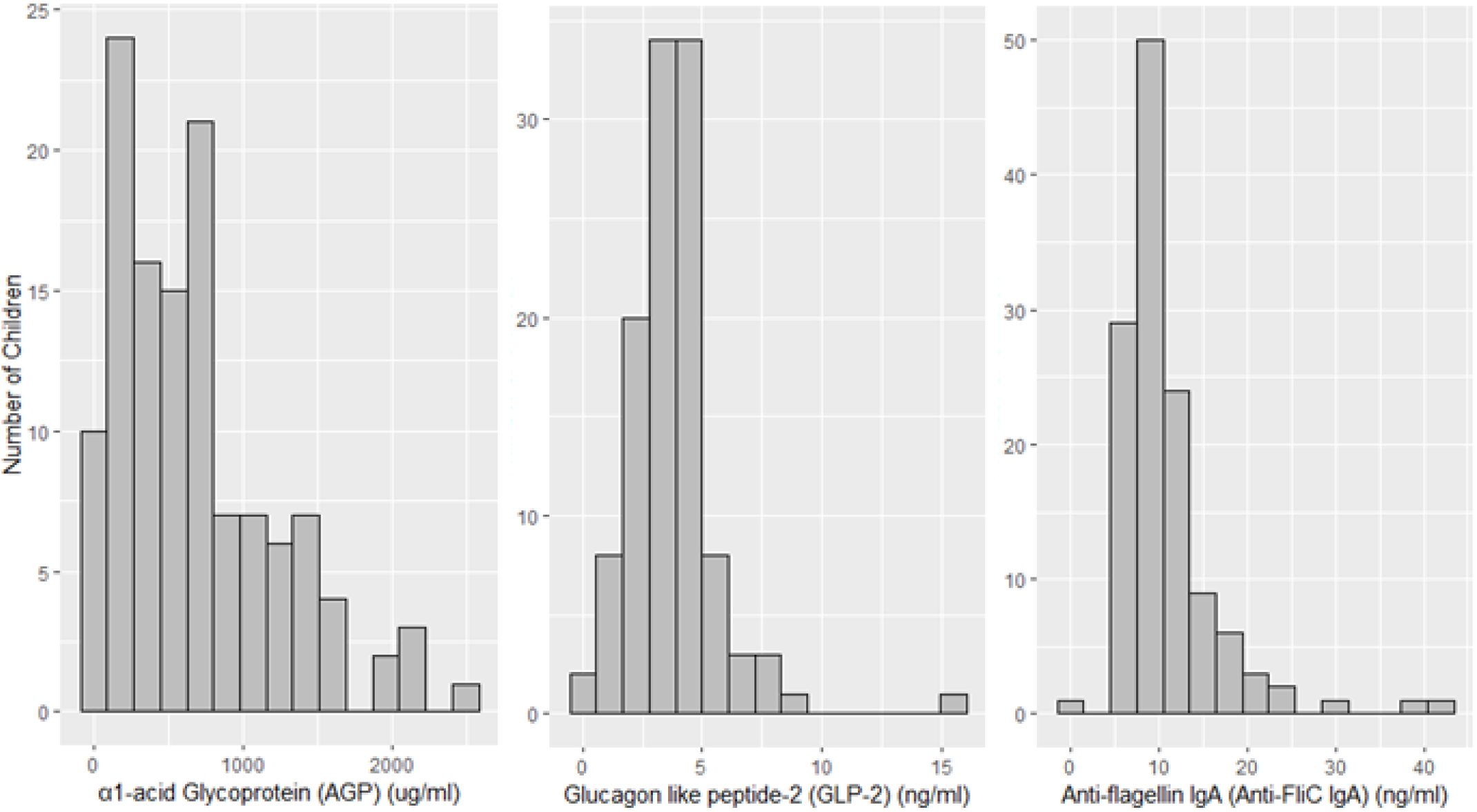
Distribution of EED biomarker concentrations: α1-acid Glycoprotein (AGP), Glucagon like peptide-2 (GLP-2), and Anti-flagellin IgA (Anti-FliC IgA)

**Table 1.**
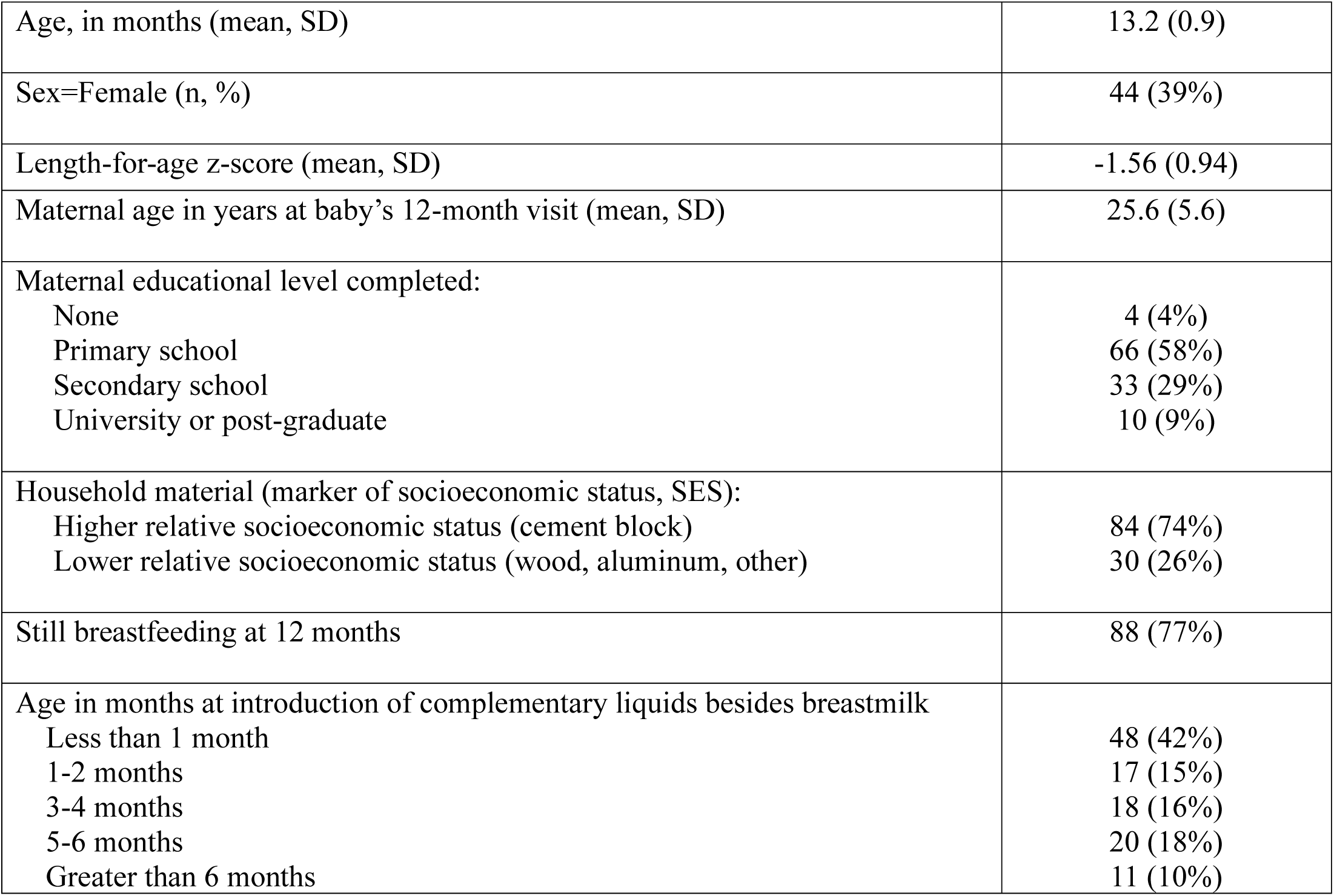
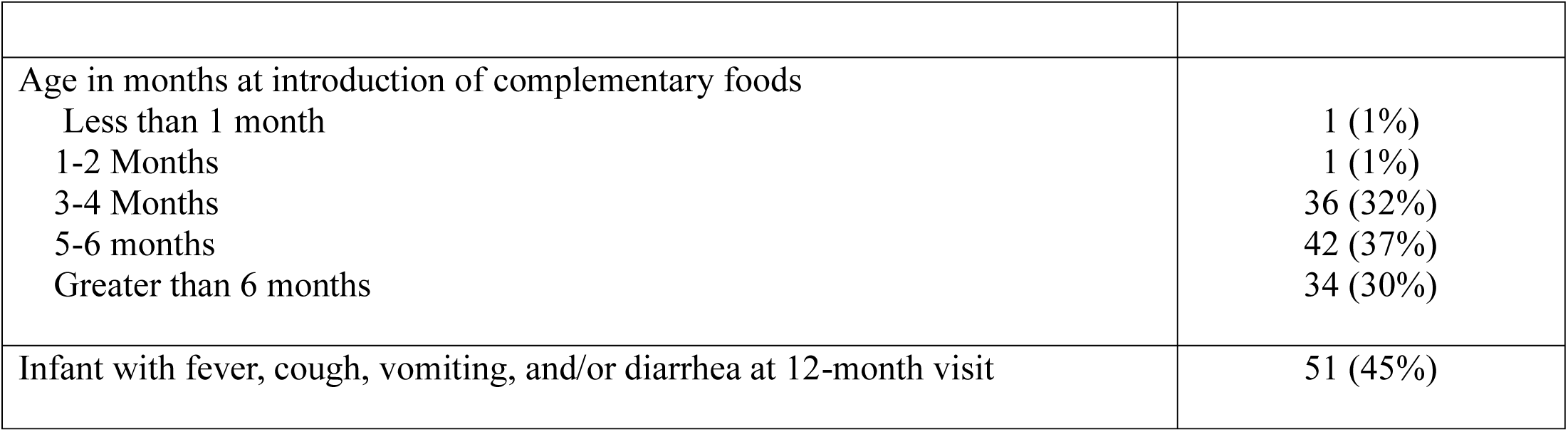
Characteristics of the 114 participating children, their mothers, and households.

|  |  |
| --- | --- |
| Age, in months (mean, SD) | 13.2 (0.9) |
| Sex=Female (n, %) | 44 (39%) |
| Length-for-age z-score (mean, SD) | -1.56 (0.94) |
| Maternal age in years at baby's 12-month visit (mean, SD) | 25.6 (5.6) |
| Maternal educational level completed: |  |
| None | 4 (4%) |
| Primary school | 66 (58%) |
| Secondary school | 33 (29%) |
| University or post-graduate | 10 (9%) |
| Household material (marker of socioeconomic status, SES): |  |
| Higher relative socioeconomic status (cement block) | 84 (74%) |
| Lower relative socioeconomic status (wood, aluminum, other) | 30 (26%) |
| Still breastfeeding at 12 months | 88 (77%) |
| Age in months at introduction of complementary liquids besides breastmilk |  |
| Less than 1 month | 48 (42%) |
| 1-2 months | 17 (15%) |
| 3-4 months | 18 (16%) |
| 5-6 months | 20 (18%) |
| Greater than 6 months | 11 (10%) |
| Age in months at introduction of complementary foods |  |
| Less than 1 month | 1 (1%) |
| 1-2 Months | 1 (1%) |
| 3-4 Months | 36 (32%) |
| 5-6 months | 42 (37%) |
| Greater than 6 months | 34 (30%) |
| Infant with fever, cough, vomiting, and/or diarrhea at 12-month visit | 51 (45%) |

**Table 2.** Correlation coefficients (and p-values) between α1-acid glycoprotein (AGP), glucagon like peptide-2 (GLP-2), and anti-flagellin IgA (Anti-FliC IgA) concentrations at 12 months of age.

|  | AGP | Anti-FliC IgA |
| --- | --- | --- |
| GLP-2 | 0.32 (0.0006) | 0.05 (0.57) |
| Anti-FliC IgA | 0.23 (0.01) | X |

In an unadjusted analysis, mean levels of the biomarkers did not differ significantly between children with and without symptoms of cough, diarrhea, vomiting, or fever, although AGP tended to be higher among the children with symptoms vs. the children without symptoms (4.11 µg/mL vs 3.47µg/mL, respectively, p=0.09). In bivariate analysis for the identification of confounders, age, maternal education, and household material were associated with both the exposure (EED biomarker concentrations) and the outcomes (LAZ and MSEL scores), while age at the introduction of complementary foods was associated with the exposure; therefore, these four variables were included in the multivariable linear regression models.

Parameter estimates (β-coefficients) and p values in the linear regression model evaluating the relationship between EED biomarkers and the outcomes are shown for all 114 children in Table 3, and among children without infectious symptoms in Table 4. Among all 114 children tested, we did not observe an association between EED biomarker concentrations and growth or neurodevelopmental outcomes. However, when we only examined children without acute infectious symptoms, we found a negative association between Anti-FliC IgA concentrations and ELC: an increase in 10 ng/L in Anti-FliC IgA concentration was associated with a decrease in the MSEL Early Learning Composite (ELC) raw score of 3.2 points.

**Table 3.**
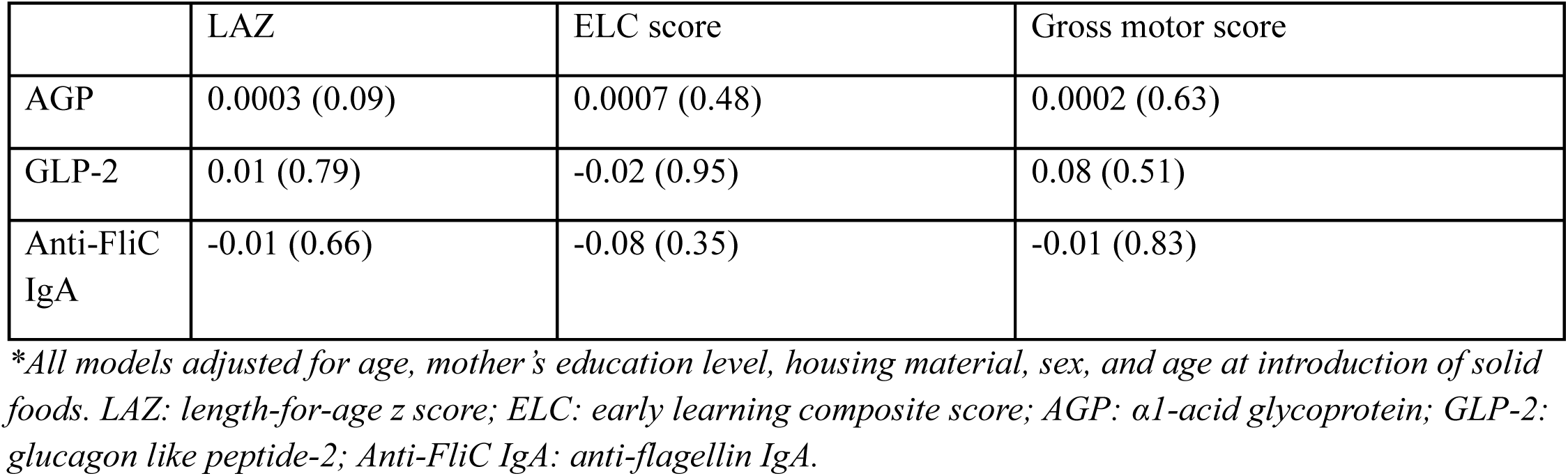
β-coefficients (p-value) for adjusted analyses* evaluating the association between biomarkers of environmental enteric dysfunction (EED) (exposures) and length-for-age Z-score (LAZ), early learning composite score (ELC), and gross motor score (outcomes) in all children at 12 months of age (N=114)

|  | LAZ | ELC score | Gross motor score |
| --- | --- | --- | --- |
| AGP | 0.0003 (0.09) | 0.0007 (0.48) | 0.0002 (0.63) |
| GLP-2 | 0.01 (0.79) | -0.02 (0.95) | 0.08 (0.51) |
| Anti-FliC<br>IgA | -0.01 (0.66) | -0.08 (0.35) | -0.01 (0.83) |

*\*All models adjusted for age, mother's education level, housing material, sex, and age at introduction of solid* *foods. LAZ: length-for-age z score; ELC: early learning composite score; AGP: α1-acid glycoprotein; GLP-2:* *glucagon like peptide-2; Anti-FliC IgA: anti-flagellin IgA.*

**Table 4:** β-coefficients (p-value) for adjusted analyses* evaluating the association between biomarkers of environmental enteric dysfunction (EED) (exposures) and length-for-age Z-score (LAZ), early learning composite score (ELC), and gross motor score (outcomes) in children without cough, diarrhea, vomiting, or diarrhea at 12 months of age (N = 63)

|  | LAZ | ELC score | Gross motor score |
| --- | --- | --- | --- |
| AGP | 0.0004 (0.099) | 0.001 (0.36) | 0.0001 (0.82) |
| GLP-2 | 0.0004 (0.99) | 0.17 (0.67) | 0.14 (0.43) |
| Anti-FliC<br>IgA | -0.01 (0.73) | **-0.32 (0.004)** | 0.00001 (0.99) |

*\*All models adjusted for age, mother's education level, housing material, sex, and age at introduction of solid* *foods. LAZ: length-for-age z score; ELC: early learning composite score; AGP: α1-acid glycoprotein; GLP-2:* *glucagon like peptide-2; Anti-FliC IgA: anti-flagellin IgA.*

## Discussion

In this exploratory analysis of infants from the lowlands of Guatemala, we did not find an association between three representative EED biomarkers and LAZ or ND at approximately 12 months of age. However, removing children with acute infectious symptoms uncovered an association between Anti-FliC IgA, a marker of increased host exposures to intestinal bacteria, and ELC score. For example, an increase in 10ng/L in Anti-FliC IgA concentration was associated with a decrease in the MSEL ELC raw score of 3.2 points, which would equate to an approximately 9-point decrease in the standard score. Assuming that the magnitude of this relationship persisted throughout early childhood, our finding suggests that a child with this degree of Anti-FliC IgA elevation persisting by age 3 could be almost 2 standard deviations in MSEL ELC score behind their peers without this degree of Anti-FliC IgA elevation. This suggests that higher host exposures to intestinal bacterial products, as may occur from increased intestinal permeability, may be a substantial risk to long-term health and development.

Although we did not observe associations between AGP and growth or ND, children in the study had higher than normal levels of AGP(43), suggesting the presence of systemic inflammation, which has been associated with poor growth in other settings. Also, children in our study had higher than normal levels of GLP-2 as compared to control children with normal growth in Zambia.(50) GLP-2 concentrations can have a paradoxical pattern—levels are higher in children with acute diarrhea (likely reflecting acute repair of the damaged intestinal epithelium), however, they are lower in children with chronic malnutrition.(50) This potentially paradoxical pattern of GLP-2 levels highlights the need to separate children with symptomatic infections or other characteristics to understand the impact of EED on long-term outcomes. Finally, we did not find strong correlations between the biomarkers representing different domains of EED. This suggests that EED is a complex of separate pathological components that may be acquired, progress independently, and may require longitudinal monitoring.

A surprisingly high proportion of children had acute infectious symptoms at the 12-month timepoint. This high burden of symptomatic infections, and likely, asymptomatic infections in this population(51) may reflect the low socioeconomic status and environmental conditions in the area and related malnutrition, which can both increase exposures and impair immune protection. It is possible that children who are sick may have transient differences in their EED biomarkers that do not reflect their baseline level, or alternately, may not perform well on the MSEL due to illness or fatigue. For this reason, we decided to perform a separate analysis that excluded children with acute infectious symptoms. As a high burden of infections have been associated with poor growth and ND, effective interventions are needed to that reduce infectious disease burden. For example, the MAL-ED multi-site cohort study showed that a higher burden of enteropathogens, specifically, those that cause mucosal disruption, were associated with elevated biomarkers of intestinal permeability and impaired growth.(44)

Higher levels of Anti-FliC IgA were related to growth faltering in young Tanzanian and Ugandan children.(26,52) A related study in Tanzania examined the association between EED biomarker levels measured at 6 months of age and ND at 15 months of age, as assessed by the Bayley Scales of Infant and Toddler Development.(28) Interestingly, Anti-FliC IgA concentrations were positively associated with ND, the opposite of what was found in our study. One notable difference is that the Tanzanian study excluded children who were stunted at baseline. Other differences in the timing of samples, tools for assessing ND outcomes, or geographic region may contribute to the differences between studies. Also, Anti-FliC IgA may be a composite measure of increased intestinal permeability as well as ability to generate protective immune responses to pathogenic bacteria. More detailed studies using a gold standard for intestinal permeability such as urinary lactulose:mannitol test (53) are needed to define a specific role for intestinal permeability defects in this population.

Similar to the Tanzanian study, we did not find an association between systemic inflammation, as measured by AGP, and ND. Another study conducted at three MAL-ED sites (Brazil, Tanzania, and South Africa) found that a higher burden of intestinal *Shigella* infection in the first two years of life was strongly associated with lower linear growth in children; systemic inflammation was associated with lower verbal fluency scores in Brazil, but not in Tanzania and South Africa.(54) These studies highlight the heterogeneity of findings across geographic locations and likely differences in host factors in the populations studied.

It is not known how elevated levels of Anti-FliC IgA may potentially influence ND. It is conceivable that children with elevated Anti-FliC IgA have impaired intestinal integrity and an increased burden of enteric infections that may result in poor absorption of micro- and macronutrients; if this occurs in infancy—a time of rapid neurological development—delays in ND may result. Interestingly, Lauer, et al, reported a strong negative association between Anti-FliC IgA and hemoglobin levels, and iron-deficiency anemia is a well-established cause of poor ND.(52) Increased intestinal permeability has also been shown to result in enteric protein loss.(55)

Our study had several limitations. First, neurodevelopmental testing of infants has some inherent limitations, as infants have a smaller repertoire of skills, a limited number of test items are administered in each developmental domain. For example, many fewer test items are administered to a 12-month-old infant in comparison to the 24-month-old child, which can make it more challenging to differentiate between children, especially those that share many risk factors related to living in poverty and adverse environmental conditions. Therefore, there may have been EED effects on ND that we were not yet able to detect due to infant age and that could be more apparent as the child gets older. Second, we measured EED biomarkers at only one timepoint. Performing longitudinal assessment of EED may more accurately reflect enteric function and its trajectory over time. Finally, we measured biomarkers representing three domains of EED. A more comprehensive panel of EED biomarkers or analysis of the gut microbiome composition may have allowed for the examination of additional domains of EED in relationship to growth and ND. (56) A strength of the study was the use of a robust performance-based measure of ND, which has been extensively validated at the research site and shown to correlate with other known ND risks, such as stunting and microcephaly.(38–40)

Future studies are warranted to further examine the link between EED and ND to guide the testing of interventions that ameliorate the EED domain(s) that may contribute to poor ND. Further, tools to more easily measure ND across field settings are needed(56,57), as growth outcomes and ND can operate independently, as was the case in our study.

In conclusion, this preliminary study examined the association between three domains of EED and ND in a population of infants at high risk for poor ND in rural SW Guatemala. Our findings suggest that intestinal permeability may play a role in poor ND outcomes. Future studies are warranted to confirm these findings in other settings, and work towards interventions to reduce the burden of poor ND in children globally.

## Data Availability

All data produced in the present study are available upon reasonable request to the authors.

## Acknowledgements

The authors would like to thank Dr. Flor M. Muñoz, Dr. Hana El Sahly, Dr. Wendy Keitel, Dr. Walla Dempsey, and Dr. Kay Tomashek for their contributions to the Parent Study as DMID and VTEU project officers and investigators. We also wish to thank the families who participated in this study, and all of the research nurses, psychologists, and other personnel from FUNSALUD who worked on the Parent Study.

